# Macrophage Infiltration and ITGB2 Expression in ESCC: A Novel Correlation

**DOI:** 10.1101/2024.05.14.24307326

**Authors:** Tao Huang, Longqian Wei, Jun Liu, Huafu Zhou

## Abstract

Esophageal squamous cell carcinoma (ESCC), one of the most prevalent and deadliest malignancies today, still presents significant limitations in the application and efficacy of immunotherapy. In this study, we comprehensively utilized high-throughput sequencing, gene chips, single-cell sequencing, and various bioinformatics analysis methods to uncover, for the first time, a positive correlation between the infiltration level of macrophages and the expression of ITGB2 in ESCC.ITGB2 is overexpressed in ESCC and holds immense potential as a prognostic marker for ESCC. As ESCC progresses, the expression of ITGB2 increases within infiltrating macrophages. We also proposed for the first time that the expression of ITGB2 in macrophages continues to increase as macrophages shift towards a pro-tumor phenotype. We found that evaluating the immune therapy response in ESCC patients using ITGB2 is feasible, and higher expression of ITGB2 correlates with increased feasibility of targeting macrophages. Additionally, we identified three miRNAs associated with aberrant expression of ITGB2, providing references for further exploration of upstream molecules of ITGB2.

## 1. INTRODUCTION

The incidence and mortality rates of esophageal cancer remain high and are on the rise^1^. Squamous cell carcinoma, as its most common histological subtype, accounts for over 70% of all global esophageal cancer cases. Currently, the 5-year survival rate for patients with esophageal squamous cell carcinoma is only around 20%^2^. Therefore, it remains a significant public health issue.

The primary treatment strategy for esophageal squamous cell carcinoma (ESCC) currently relies on comprehensive treatment, with surgery as the main approach. However, the overall treatment outcomes are not satisfactory^3^. With continuous advancements in tumor research, immunotherapy, as a novel treatment modality, has opened new avenues for ESCC treatment, following the success achieved in tumors such as lung cancer, renal cancer, and melanoma.^4^. Some clinical trials have also achieved certain results, such as Checkmate-577^5^, Keynote-97^6^, and Keynote-590^7^.

But, several factors hinder the further application of immunotherapy in ESCC. Studies have reported that patients initially responsive to immunotherapy tend to develop Acquired Resistance (AR) after receiving two or more courses of treatment^8^. Additionally, most current therapies require Programmed Cell Death 1 Ligand 1 (PDL1) positivity. Due to tumor heterogeneity, some patients present as PDL1 negative, rendering them unable to benefit from the numerous current regimens^9^. It is well known that Programmed Cell Death 1 (PD1) / PDL1 is associated with T cell activation. However, the Tumor Microenvironment (TME) is complex, comprising various immune cells. The infiltration of other immune cells in tumors also affects tumor development and patient prognosis^10^. Thus, PD1/PDL1 is just a starting point, and further exploration of more tumor immune-related biomarkers, targets, and molecular mechanisms is still needed in the future.

Today, with the development and maturation of high-throughput sequencing, gene chips, single-cell sequencing, and related bioinformatics analysis technologies, we have been provided with a wealth of tumor-related genomic information, offering unprecedented opportunities to explore biomarkers, targets, and molecular mechanisms related to tumor immunity.

In this study, we will comprehensively employ high-throughput sequencing, gene chips, single-cell sequencing, and various bioinformatics analysis methods to explore pivotal genes involved in ESCC immune infiltration. We will delve into the relationship between these pivotal genes and the development of immune cells, as well as its potential applications in immunotherapy.

## 2. MATERIALS AND METHODS

### 2.1 Data Source and Preprocessing

The gene chip data were obtained from four series in the GEO(Gene Expression Omnibus) database: GSE161533, GSE23400,GSE66274, and GSE67268. To enhance data reliability, all chip data underwent cleaning, standardization, inspection, and batch effect removal. Subsequently, probe annotation and gene ID conversion were performed.

The high-throughput sequencing data were sourced from The Cancer Genome Atlas (TCGA) and the Genotype-Tissue Expression (GTEx) Database. To ensure data reliability, we utilized the TCGA original count data from UCSC Xena and systematically checked each TCGA sample using the “Merged Sample Quality Annotations” file from the TCGA website, removing samples with poor quality. Only high-quality samples with stage “A” were retained. Moreover, for esophageal squamous cell carcinoma samples, we selected samples with “Sample Type Codes” of “01”, “02”, “05”, or “08”. For normal tissue samples, we only chose samples with “Sample Type Codes” of “11”. Considering the different sources of data from TCGA and GTEx, direct merging analysis could introduce significant bias and affect the accuracy of results. Therefore, we utilized the “UCSC Toil RNAseq Recompute Compendium” ^11^data from UCSC Xena for merged analysis to minimize bias arising from different data sources. For the original count data, batch effect correction was performed using the “batch_number” provided by UCSC Xena. Since the data from “UCSC Toil RNA-seq Recompute Compendium” did not provide batch effect reference, we explored and removed batch effects using the R package “RUVSeq(v1.32.0)”.

The single-cell sequencing data were obtained from the GEO database under accession number GSE199619. We utilized the “GSE199619_ELN.integrated.rds.gz” file. To minimize bias, we selected ESCC samples that underwent surgical resection or endoscopic resection without neoadjuvant chemotherapy (NACT) (Supplementary Table 2).. Summary information for all data is provided in Supplementary Table 1.

### 2.2 Immunoinfiltration Analysis and WGCNA

We conducted immunoinfiltration analysis using ImmuCellAI. The preprocessed gene expression matrices from GSE161533 and GSE23400 were imported into ImmuCellAI, and the “Analysis” function was utilized to quantify the infiltration of various immune cells in tumor samples.

Subsequently, based on the gene expression matrices and corresponding immunoinfiltration analysis results, we constructed a weighted gene co-expression network using the R package “WGCNA (1.71)” and screened for gene modules associated with various immune cell infiltrations. Specifically, we first selected 10,000 genes with large standard deviations and identified outlier samples through sample clustering, followed by their removal. Then, we chose an appropriate soft threshold to construct the scale-free network and TOM matrix. Next, we calculated the correlation between gene modules and immune cell infiltration using the “moduleTrait” function. If multiple associations were found between immune cells and gene modules, we applied two criteria for selection: (1) selecting immune cells with higher average infiltration abundance and (2) conducting correlation tests with P-values less than 0.01. Subsequently, we obtained immune infiltration-related genes by intersecting the gene modules obtained from GSE161533 and those obtained from GSE23400.

### 2.3 Enrichment Analysis and Core Gene Selection

We annotated the biological significance of intersecting genes using the R package “clusterProfiler (4.6.0)” to indirectly demonstrate the correlation between these genes and infiltrating immune cells. This annotation involved two enrichment analysis methods: Gene Ontology (GO) and Kyoto Encyclopedia of Genes (KEGG).

Subsequently, we imported the intersecting genes into the STRING database to construct a Protein-Protein Interaction (PPI) network. Associations with the “combined score” less than 0.7 were excluded to obtain a reliable PPI network. The network was then imported into Cytoscape (3.9.2), and key genes in the network were calculated using the “MCC” method in the cytoHubba tool. Finally, the top ten ranked genes were selected as the core genes of the PPI network.

### 2.4 Gene Expression Differential Analysis and Survival Analysis

For chip data, we used the R package “limma (3.54.2)” to perform paired comparisons between ESCC samples and normal esophageal tissue samples from the GSE161533 and GSE23400 datasets to analyze gene expression differences. For high-throughput sequencing data, we extracted RNA counts data for ESCC and normal esophageal tissue from the “UCSC Toil RNAseq Recompute Compendium” dataset and conducted differential analysis using the R package “DESeq2 (1.38.1)”.

As for survival analysis, we obtained data from TCGA. Utilizing the R package “survminer (0.4.9)”, we determined the optimal cutoff point and then assessed whether there were differences in overall survival (OS), disease-specific survival (DSS), and progression-free interval (PFI) between patients with high expression of core genes and those with low expression using the R package “survival (3.4.0)”.

Finally, we selected the core genes from the PPI network that were significant in both expression differential analysis and survival analysis as target genes. The immune cells corresponding to the gene module where the target genes were located were identified as target cells.

### 2.5 Validation of the correlation between target genes and target cells

For chip data, we extracted the infiltration abundance of target cells from the previous ImmuCellAI results, and then performed correlation analysis between the expression values of target genes extracted from the gene expression matrix and the infiltration abundance of target cells. For high-throughput sequencing data, we first downloaded the immune infiltration data for esophageal cancer from the ImmuCellAI website, extracted the infiltration abundance of target cells in ESCC, and then converted the count data of ESCC to TPM data. Subsequently, we conducted correlation analysis between the TPM data of target genes and the infiltration abundance of target cells.

Additionally, to enhance the reliability of the results, we further utilized six different algorithms (TIMER, CIBERSORT, CIBERSORT.ABS, QUANTISEQ, XCELL, EPIC) from the TIMER2 website to calculate the immune infiltration abundance of target cells originating from ESCC samples in TCGA. Subsequently, we performed correlation analysis between the results of these additional algorithms and the TPM data of target genes.

### 2.6 Validation of the Protein Expression of the Target Gene

We conducted Western blot (WB) and immunohistochemistry (IHC) experiments to examine the protein expression of the target gene. We collected a total of 8 pairs of biological samples from 4 patients, including 8 ESCC tissues and 8 adjacent normal esophageal tissues. Four pairs of samples were used for WB experiments, and the other four pairs were used for IHC experiments. All patients were unrelated Asians who were hospitalized and treated at the First Affiliated Hospital of Guangxi Medical University (Guangxi Province, China). ESCC was confirmed by histopathological examination of tissue obtained from surgical resection of the tumor or biopsy. None of the patients received chemotherapy or radiotherapy before tumor resection. Biological samples were collected immediately after tumor resection and analyzed according to the specified procedures (Supplementary Materials 1-2).

### 2.7 Mining of Single-cell RNA Data

First, we compared the expression differences of the target genes in different cells using the R package “Seurat(4.3.0)”. Subsequently, we explored whether the expression of the target genes was related to the differentiation and development of target cells using the R packages “IOBR(0.99.9)” and “Monocle(2.26.0)”. We also analyzed whether different tumor stages affect the expression of target genes in the target cells.

### 2.8 Immunotherapy Response

We selected TME signature gene sets related to immunotherapy response based on relevant literature and gene sets collected by R package “IOBR (0.99.9)”. Utilizing the ESCC data from the GSE23400 dataset and the TPM data from TCGA, we quantified these TME signatures using the “ssGSEA” algorithm in the R package “IOBR(0.99.9)”. Subsequently, correlation analysis was conducted to understand the association between the target genes and these signatures.

### 2.9 Correlation between Target Genes and Known Targets of Target Cells

We identified the primary known targets of the target cells through literature review and correlated these targets with the target genes using gene expression data from GSE23400 and TPM data from TCGA, conducting correlation analysis to assess the association.

### 2.10 Predicting Upstream miRNAs

Initially, we utilized miRWork to predict miRNAs that may potentially bind to the target genes. Subsequently, we filtered out miRNAs with a binding probability greater than 0.9. Differential expression analysis was conducted using data from GSE66274 and GSE67268 to identify miRNAs exhibiting differential expression between ESCC and esophageal normal tissues. Following this, we further refined the selection by filtering for miRNAs with an adjusted p-value less than 0.01 and a fold change (log_2_FC) less than −2. Finally, we intersected the miRNAs obtained from GSE66274, those obtained from GSE67268, and those selected from miRWork to obtain the final miRNAs.

## 3 RESULTS

### 3.1 Immune Infiltration Status and WGCNA in ESCC

The immune cell profiling using ImmuneCellAI revealed a significant presence of dendritic cells, B cells, macrophages, NK cells, and other immune cell types within ESCC (Figure 1A). (Click on the figure to view the image) However, the abundance of other immune cell types is comparatively lower. Consequently, these ten immune cell types were included for subsequent analysis.

**FIGURE 1.**
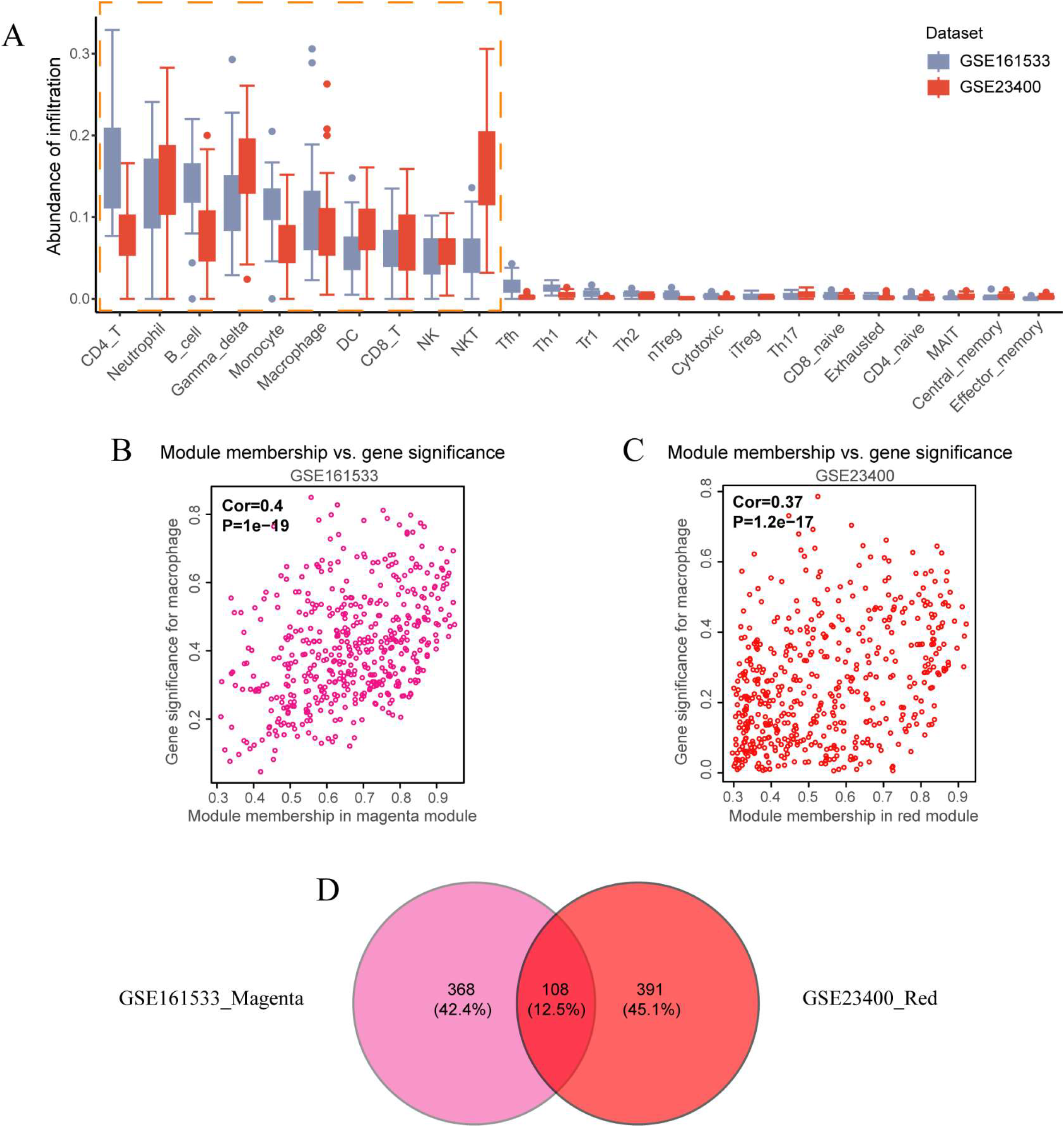
ImmuCellAI and WGCNA results. **A**. infiltration abundance of different immune cells in ESCC (Derived from ImmuneCellAI). **B**. The correlation between the magenta module and macrophages in GSE161533. **C**. The correlation between the red module and macrophages in GSE23400. **D**. Venn diagram showing the intersection between the magenta module in GSE161533 and the red module in GSE23400. (Click on A, B, C, or D to return to the corresponding paragraph)

For GSE161533, we set the “cutHeight” parameter of WGCNA to 95, removing 3 outlier samples (GSM4909616, GSM4909627, GSM4909622), and then selected a soft threshold of 6 to fit the optimal scale. At this point, R_2_ = 0.901 (Supplementary Figure 1A), and the average network connectivity was 33.5 (Supplementary Figure 1C). After constructing the weighted gene co-expression network and merging modules with correlation coefficients greater than 0.75, all genes were assigned to 16 different colored gene modules (Supplementary Figure 1E). Through correlation analysis, we found multiple associations between gene modules and immune cell infiltration (Supplementary Figure 2A), such as a significant correlation between the magenta module and macrophage infiltration (P=0.001), and between the yellow module and NK cell infiltration (P=0.001).

For GSE23400, we set the cutHeight to 65, removing 4 outlier samples (GSM573938, GSM573951, GSM573901, GSM573906), and then selected a soft threshold of 5 to fit the optimal scale. At this point, R_2_= 0.851 (Supplementary Figure 1B), and the average network connectivity was 26.9 (Supplementary Figure 1D). After constructing the weighted gene co-expression network and merging modules with correlation coefficients greater than 0.75, all genes were assigned to 12 different colored gene modules (Supplementary Figure 1F). Through correlation analysis, we also found multiple associations between gene modules and immune cell infiltration (Supplementary Figure 2B), such as a significant correlation between the yellow module and neutrophil infiltration (P=0.001), and between the red module and macrophage infiltration (P=0.002).

Therefore, we screened using the two predefined filtering criteria and determined one gene module from each of the two sets of WGCNA results for further analysis. The magenta module was identified for GSE161533, and the red module for GSE23400 (Supplementary Figure 2). The corresponding immune cell was macrophages. Subsequent analysis revealed a significant positive correlation between module membership and gene significance for both groups (Figure 1B-C). Thus, taking the intersection of the two gene modules yielded 108 genes associated with macrophage infiltration (Figure 1D).

### 3.2 Enrichment Analysis and PPI Network Screening

The outcomes of KEGG analysis suggested that the intersecting genes were linked to biological processes like “Phagosome,” “Osteoclast differentiation,” “Lysosome,” “NF-kappa B signaling pathway,” and “Antigen processing and presentation” (Supplementary Figure 3A). GO analysis results indicated that the intersecting genes were associated with biological characteristics including “macrophage activation,” “phagocytosis,” “cell activation involved in immune response,” “myeloid leukocyte activation,” “myeloid leukocyte migration,” “ficolin-1-rich granule,” “ficolin-1-rich granule lumen,” “phagocytic cup,” “endocytic vesicle membrane,” “integrin complex,” “mannose binding,” “immunoglobulin binding,” “immune receptor activity,” “pattern recognition receptor activity,” and “complement binding” (Supplementary Figure 3B). These findings further emphasize the probable correlation between the expression of intersecting genes and macrophage infiltration in ESCC.

As shown in Supplementary Figure 4A, using the STRING database, we obtained the protein-protein interaction (PPI) network of the relevant genes. Associations with a “combined score” less than 0.7 were excluded (Supplementary Figure 4B), and using the “MCC” method of the cytoHubba tool for calculation, we ultimately identified ten core genes in the network (Supplementary Figure 4C). Ranked from high to low based on their scores, they are: CSF1R, ITGB2, ITGAM, TYROBP, FCGR2A, CD14, FCER1G, ITGAX, TLR4, and C1QB.

### 3.3 Gene Expression Differential Analysis and Survival Analysis

Gene expression differential analysis and survival analysis revealed significance in ITGB2, ranked second among these ten core genes, in both analyses (Figure 2, Supplementary Tables 8-9). Results from three different datasets consistently showed higher expression of ITGB2 in ESCC compared to esophageal normal tissues (all P-values < 0.05, Figures 2A-C). Patients with high expression of ITGB2 in ESCC had shorter OS, DSS, PFI times compared to those with low expression of ITGB2 (all P-values < 0.05, Figures 2D-F).

**FIGURE 2.**
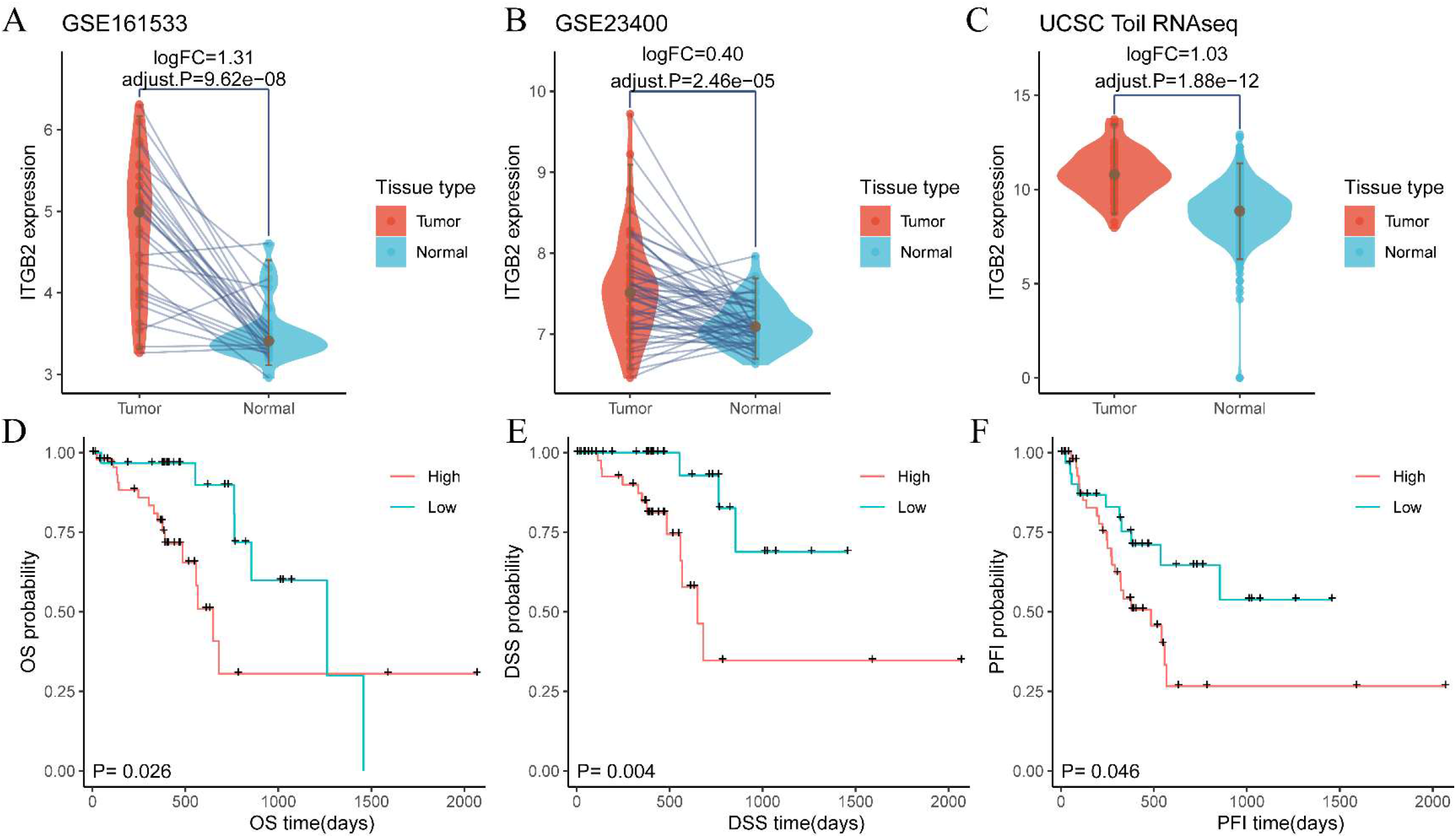
Expression Differential Analysis and Survival Analysis Results for ITGB2. **A-C**. Expression Differential Analysis Results. **A**. GSE161533. **B**. GSE23400. **C**. UCSC Toil RNA. **D-F**. Kaplan-Meier Curves for Different Survival Metrics. **D**. overall survival (OS). **E**. disease-specific survival (DSS). **F**. progression-free interval (PFI).

Furthermore, both immunohistochemistry (IHC) and Western blot (WB) results demonstrated significant upregulation of ITGB2 protein in ESCC compared to esophageal normal tissues (all P-values < 0.05, Figure 3).

**FIGURE 3.**
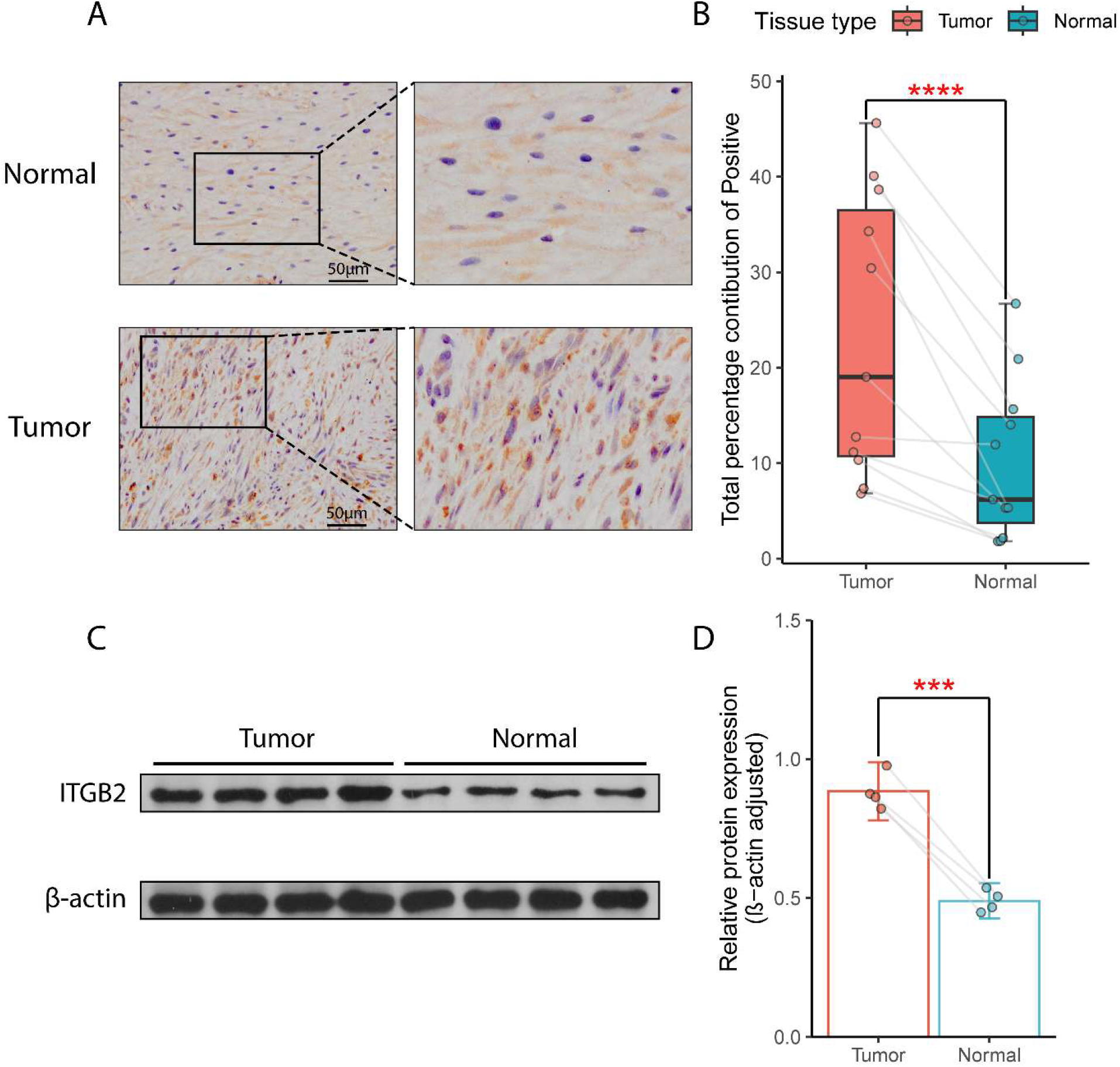
**A**. Representative IHC images of tumor samples and normal tissue samples. **B**. Differential analysis of IHC based on quantification using ImageJ. **C**. WB results of tumor samples and normal tissue samples. **D**. Differential analysis of WB based on quantification using ImageJ.

These findings suggest that ITGB2 is a promising and worthy gene for further investigation. The overexpression of ITGB2 is likely associated with the occurrence and progression of ESCC, and this correlation may be closely related to macrophage infiltration. Therefore, we identified ITGB2 as the target gene for this study. Correspondingly, macrophages were identified as our target cells.

### 3.4 Examination of the Association between ITGB2 and Macrophage Infiltration

In all three different datasets, the expression of ITGB2 was significantly positively correlated with macrophage infiltration (P < 0.05, Figure 4 (A, C, E)). In the GSE161533 dataset, the Spearman correlation coefficient (rho) was 0.7 with a P-value of 4e-05 (Figure 4B); in the GSE23400 dataset, the Spearman correlation coefficient was 0.39 with a P-value of 3.7e-03 (Figure 4D); in the TCGA dataset, the Spearman correlation coefficient was 0.64 with a P-value of 1.9e−10 (Figure 4E). Additionally, results from multiple additional methods also indicated a significant positive correlation between ITGB2 expression and ESCC macrophage infiltration (Supplementary Figure 5).

**FIGURE 4.**
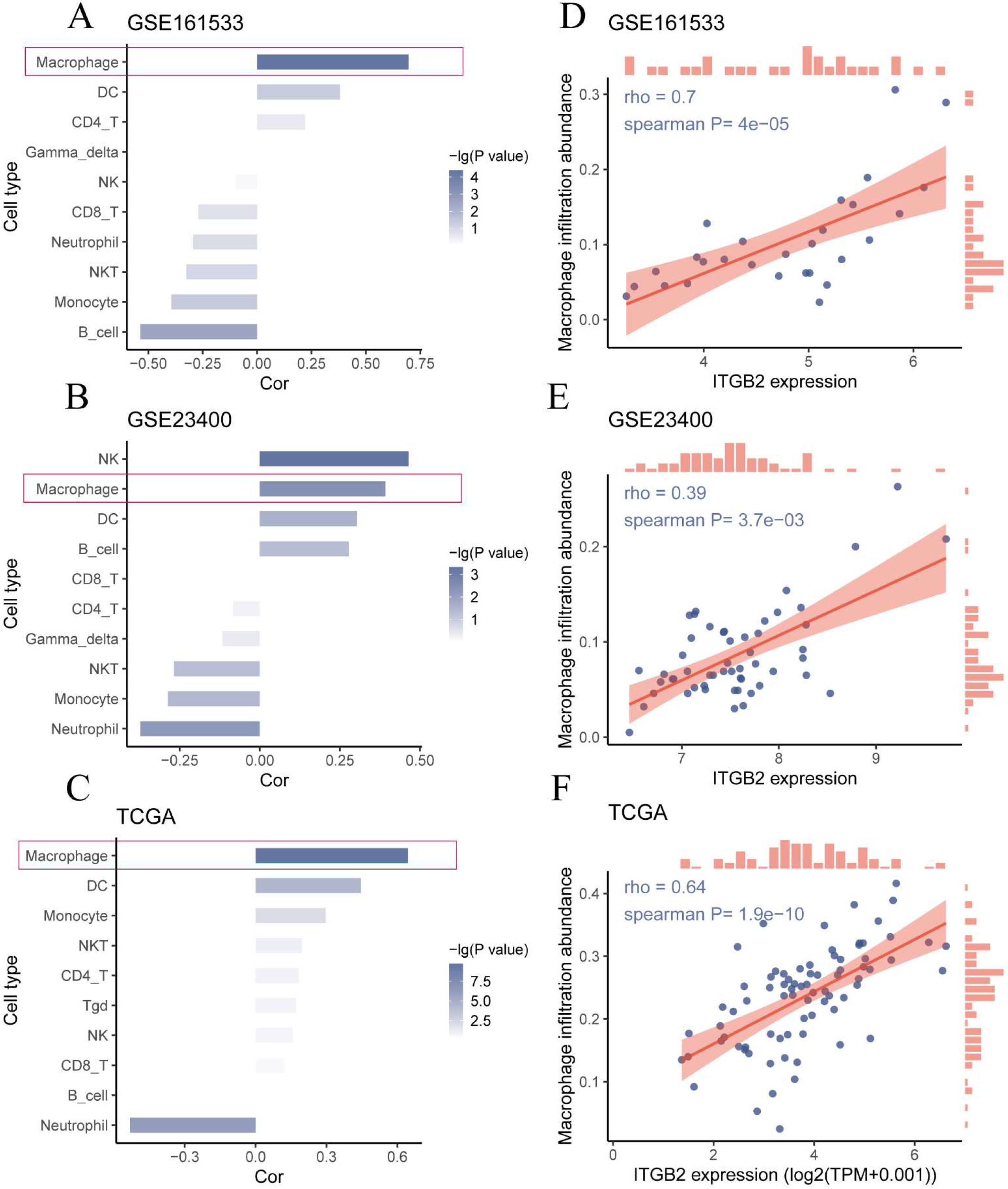
Correlation between ITGB2 expression and immune cell infiltration. **A-C**. Correlation of ITGB2 expression with various immune cells across different datasets. **A**. GSE161533. **B**. GSE23400. **C**. TCGA. **D-F**. Correlation of ITGB2 expression with macrophage infiltration across different datasets. **D**. GSE161533. **E**. GSE23400. **F**. TCGA.

### 3.5 Parsing Single-Cell RNA Data

The single-cell sequencing samples comprised 20,793 cells (Supplementary Figure 6A). We classified cells using DCN, PDPN, CD2, CD3D, ITGAX, CD68, CD79A, KRT5 EPCAM, and TPSB2 as markers, as detailed in Supplementary Figure 6B. Subsequently, we isolated macrophages from myeloid cells using CD14 as a marker (Supplementary Figure 6C-D), obtaining a total of 3491 macrophages. We observed that ITGB2 expression in macrophages was higher than in other cell types (Figure 5A, Supplementary Table 11).

**FIGURE 5.**
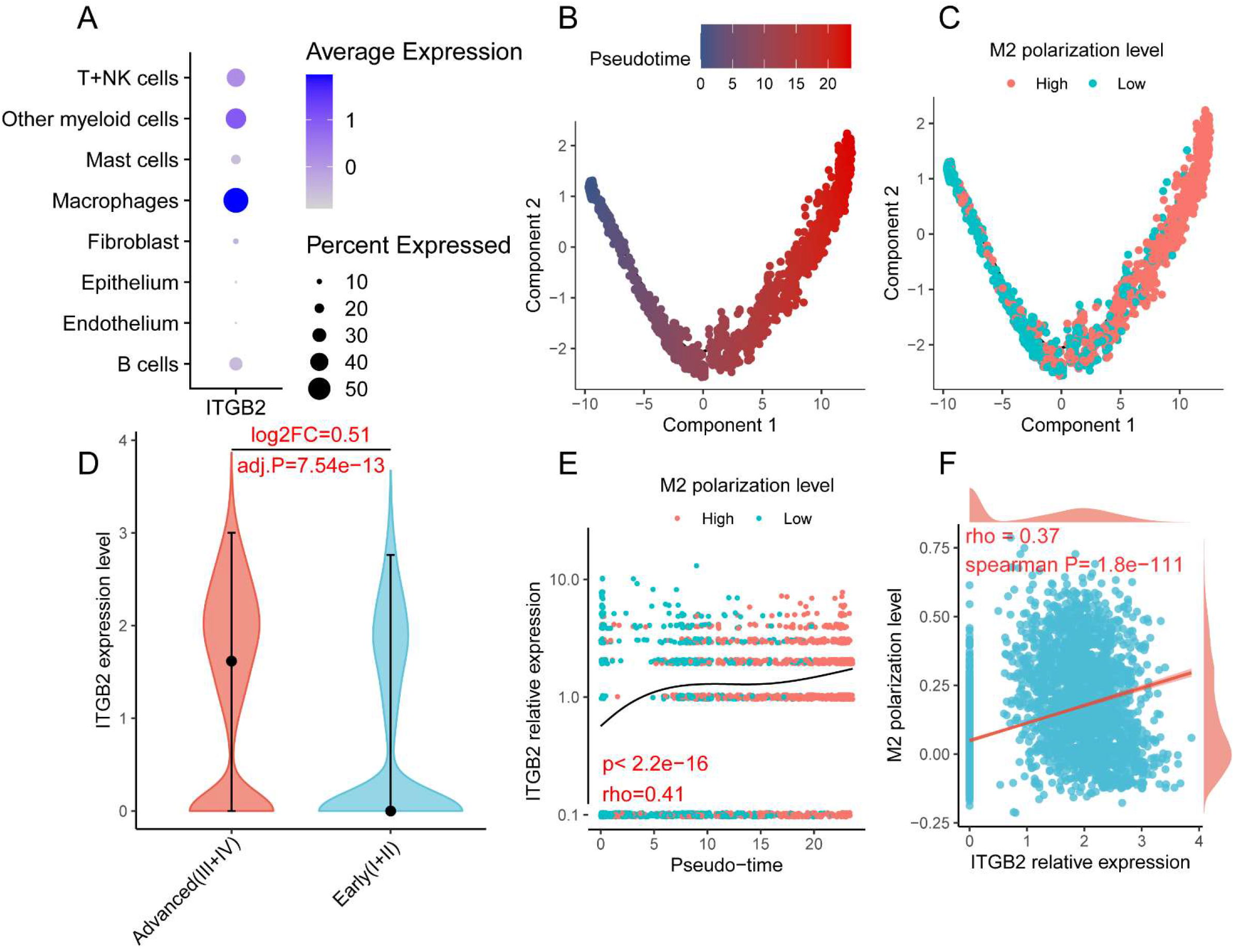
The analysis results of single-cell sequencing data. **A**. Expression profile of ITGB2 across different cell types. **B**. Pseudotime analysis results of macrophages. **C**.M2 polarization levels of macrophages at different time points. **D**. Differential expression levels of ITGB2 in macrophages between early and late stage ESCC. **E**. Correlation between macrophage development and intracellular expression levels of ITGB2. **F**. Correlation between intracellular expression levels of ITGB2 and M2 polarization levels of macrophages.

We then performed pseudotime analysis on the macrophages. Following the widely accepted cancer immune editing theory^12^, we assessed whether pseudotime conformed to an objective pattern; namely, macrophages in the tumor microenvironment would progressively acquire pro-tumor characteristics. Over time, macrophages would exhibit increasingly significant M2 features. We quantified the strength of M2 features in each macrophage using the “ssGSEA” algorithm in the R package “IOBR.” Details of the M2 feature gene set are provided in Supplementary Table 12.

The results confirm the reliability of our pseudotime analysis. As time progresses, the M2 characteristics of macrophages become more prominent (Figure 5B-C). We found a positive correlation between the expression of ITGB2 within macrophages and pseudotime, as well as M2 characteristics (all P-values < 0.05, Figure 5E-F). Further analysis revealed that in the late-stage microenvironment of ESCC, the expression of ITGB2 within macrophages is even higher (adjusted P-value = 0.54e-43, Figure 5D).

### 3.6 Immunotherapy Response

By reviewing relevant literature and referring to various gene sets collected by “IOBR”, we screened 16 gene sets associated with tumor immunotherapy response (Supplementary Table 13). Through correlation analysis, we found that the expression of ITGB2 is positively correlated with these 16 TME features related to immunotherapy response, in both the GSE23400 and TCGA datasets (all P-values < 0.05, correlation coefficients > 0, Figure 6, Supplementary Table 14).

**FIGURE 6.**
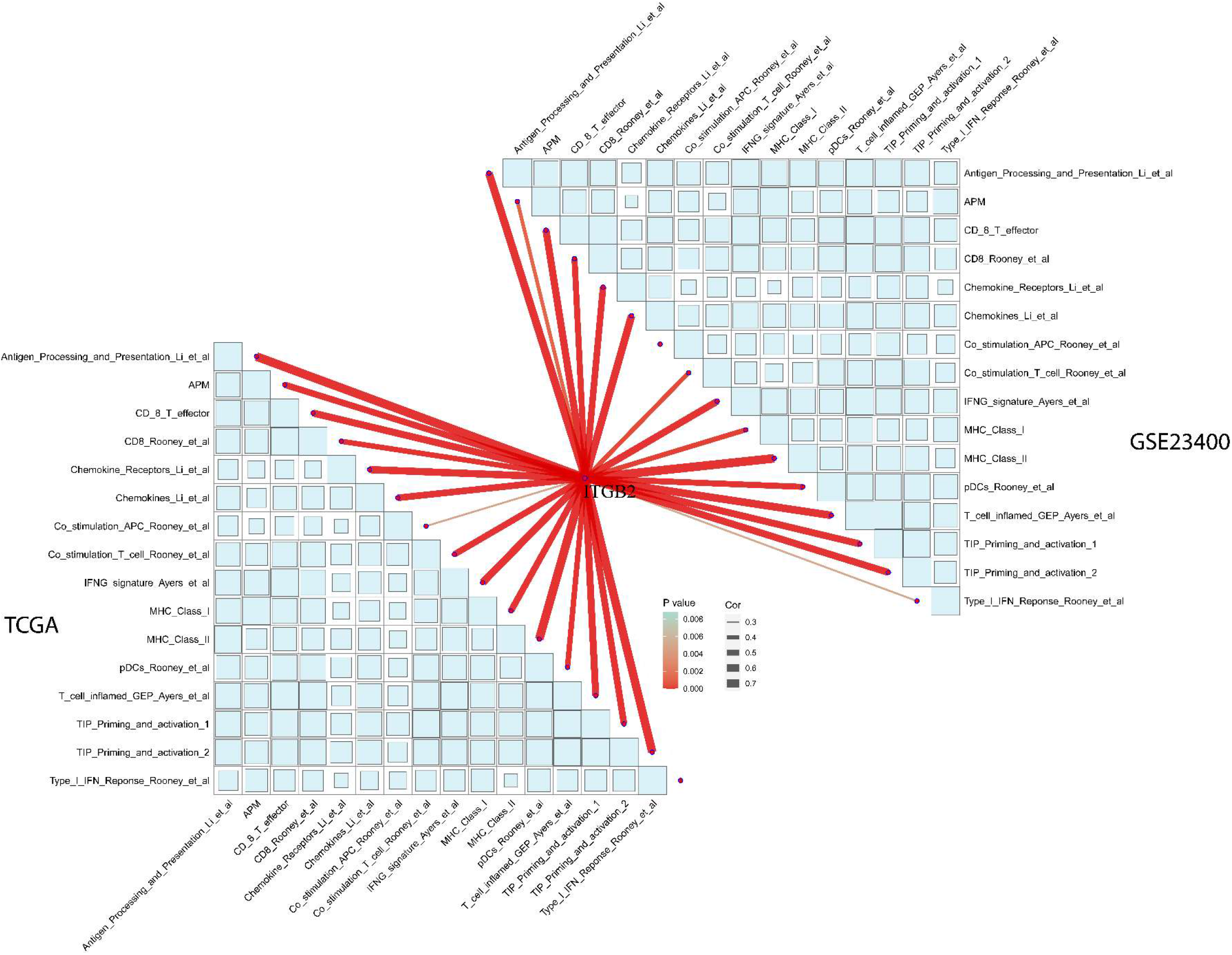
The correlation between ITGB2 expression in ESCC and 16 immune therapeutic response-related TME signatures.

### 3.7 Correlation of ITGB2 with Reported Targets of Macrophages

Through a review of relevant literature, we identified ten major known targets of macrophages, namely CCL2, CCR5, CSF1R, CD47, CD40, TLR3, TLR7, and TREM2. Clinical trials related to these targets are currently underway, with some having achieved certain results (Supplementary Table 15). We found that in both the GSE23400 and TCGA datasets, ITGB2 is significantly positively correlated with these targets (P-values all < 0.05, and correlation coefficients all > 0) (Figure 7).

**FIGURE 7.**
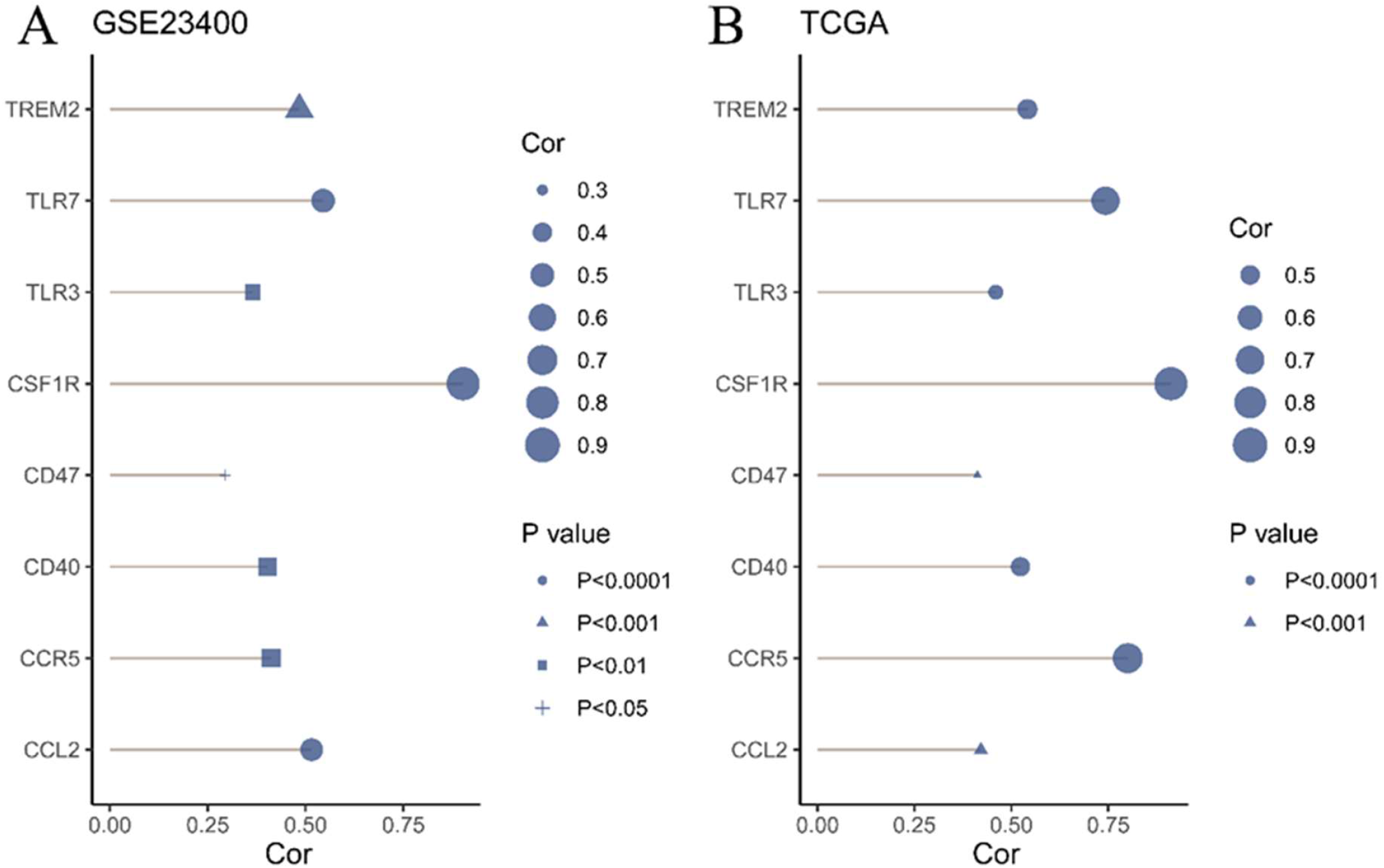
The correlation between ITGB2 and known major targets of macrophages in different datasets. **A**. GSE23400. **B**. TCGA

### 3.8 Upstream miRNA

Utilizing miRWork, we identified 2796 miRNAs with a binding probability greater than 0.9 to ITGB2 (Figure 8A). Through the aforementioned screening methods, we ultimately identified three key miRNAs (hsa-miR-18a, hsa-miR-196a, hsa-miR-21, Figure 8B). In the GSE66274 dataset, compared to esophageal normal tissues, these three key miRNAs were downregulated in ESCC (P < 0.001, Figure 8C). Similar results were observed in the GSE67268 dataset (P < 0.0001, Figure 8D).

**FIGURE 8.**
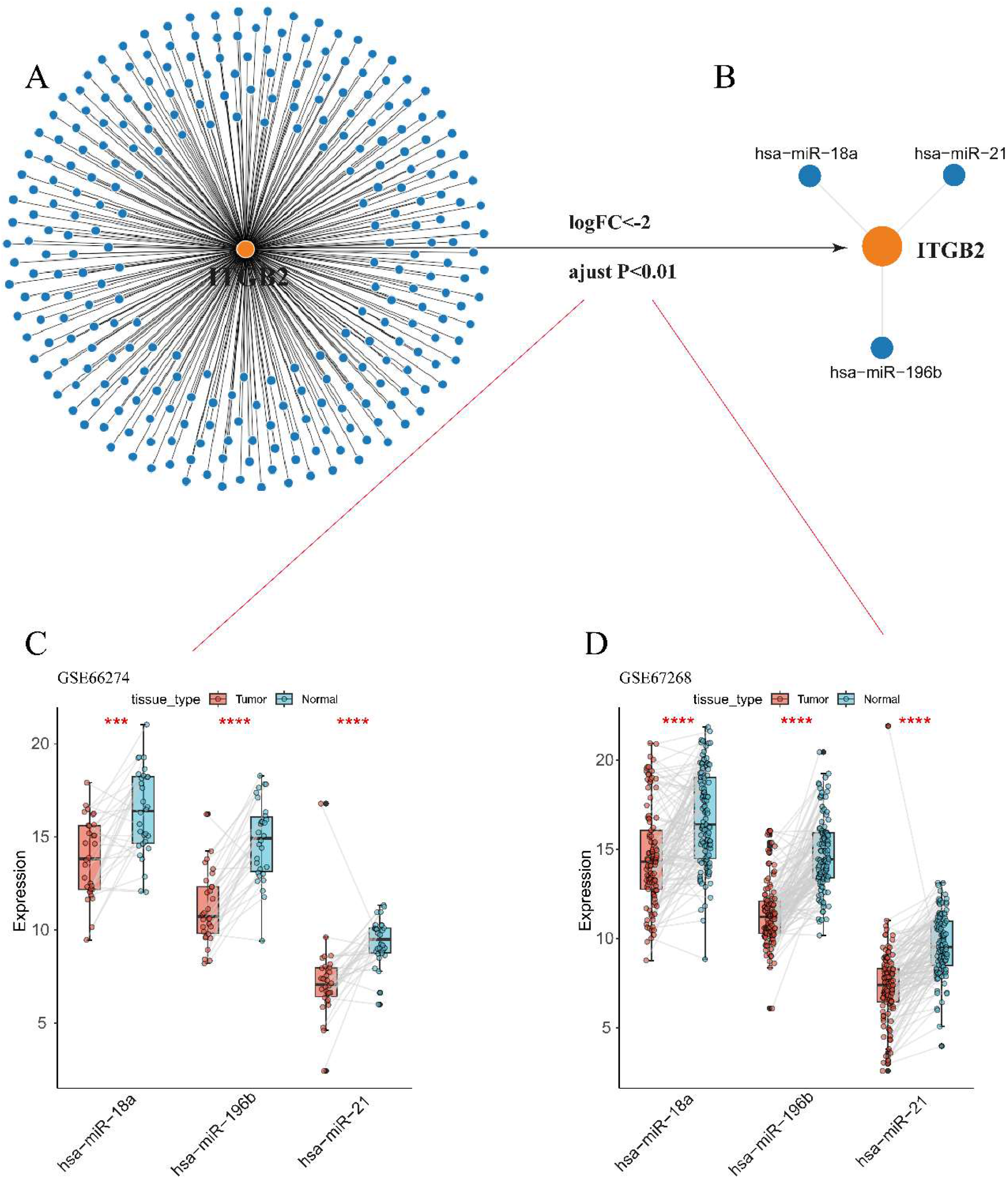
Analysis results of the miRNA-ITGB2 interaction. **A**. miRNAs with a binding probability greater than 0.9 in miRWork. **B**. Key miRNAs obtained through screening. **C**. Differential expression of key miRNAs between ESCC and normal esophageal tissues in GSE66274. **D**. Differential expression of key miRNAs between ESCC and normal esophageal tissues in GSE67268.

## 4. DISCUSSION

First and foremost, it is undeniable that there are some differences between the quantification of ESCC immune infiltration by ImmuCellAI and the cell clustering results from scRNA, owing to technical disparities. However, it is equally undeniable that both techniques’ outcomes indicate a rich infiltration of macrophages in ESCC. Presently, due to the heterogeneity and complexity of the TME^13^, the application and efficacy of immunotherapy in ESCC remain limited. Nevertheless, research has shown that tumor tissue-infiltrating macrophages can influence tumor development through multiple mechanisms, underscoring their significant research and potential clinical utility^13,14^.

ITGB2 (integrin subunit beta 2) is the beta subunit of β2 integrins. ITGB2 can form four different types of β2 integrins with four different alpha subunits (CD11a, CD11b, CD11c, CD11d). The functions of the four types of β2 integrins are mostly similar^15^. Initially discovered to be expressed in leukocytes, ITGB2 promotes leukocyte adhesion to endothelial cells, leading to extravasation^16^. Nowadays, with the continuous deepening of research, ITGB2 has been found to be associated with some cancers. For example, Paierhati et al. found that the expression level of ITGB2 in TNBC (Triple-Negative Breast Cancer) is significantly higher than in normal breast tissue, and high expression of ITGB2 in TNBC affects patient prognosis^17^. Xu et al. suggested that ITGB2 could serve as a novel prognostic factor for clinical outcomes and immune therapy response in gliomas, and it could also be a target for immune therapy in glioma patients^18^. Li et al.’s study indicated that ITGB2 is overexpressed in ovarian cancer tissues and can serve as a prognostic marker for ovarian cancer^19^. These findings underscore the value of ITGB2 in oncological research and its potential for clinical applications. However, there is still limited research on the correlation between ITGB2 and tumor-associated macrophage infiltration. We have yet to identify literature documenting the association between ITGB2 and macrophage infiltration in ESCC.

To our knowledge, we are the first to confirm the correlation between macrophage infiltration in ESCC and the expression of ITGB2. In this study, we comprehensively employed various bioinformatics approaches to identify ITGB2 as a gene closely associated with macrophage infiltration in ESCC. Through data from different sources and multiple methods, we have confirmed the positive correlation between macrophage infiltration and ITGB2 expression in ESCC. Moreover, during the screening process and further analysis, we obtained additional meaningful results related to it.

Firstly, the results from three distinct datasets consistently demonstrate the overexpression of ITGB2 in ESCC. Additionally, IHC and WB results indicate higher protein levels of ITGB2 in ESCC compared to normal esophageal tissues. Furthermore, ESCC patients with high ITGB2 expression exhibit shorter overall survival (OS), disease-specific survival (DSS), and progression-free interval (PFI) compared to those with low ITGB2 expression. These findings strongly support the reliability of ITGB2 overexpression in ESCC and its significant potential as a prognostic biomarker.

Secondly, through the exploration of single-cell sequencing data, we discovered that within the microenvironment of ESCC, the expression of ITGB2 in macrophages is significantly higher compared to other cells. It is well-known that under the influence of the tumor microenvironment, macrophages continually evolve towards a tumor-promoting phenotype^12^. Through pseudotime analysis, we observed a positive correlation between ITGB2 expression and the development of macrophages, as well as M2 characteristics. This indicates that the expression of ITGB2 within macrophages increases progressively as they evolve towards a tumor-promoting phenotype. This undoubtedly represents a novel finding, offering new insights for targeted immunotherapy directed at macrophages. Additionally, we observed that in the microenvironment of advanced-stage ESCC, the expression of ITGB2 in macrophages is higher compared to early-stage ESCC. This finding also suggests that with ESCC progression, the expression of ITGB2 within macrophages increases.

Thirdly, we further explored the feasibility of using ITGB2 to evaluate immune therapy response. We found that ITGB2 is positively correlated with 16 TME features related to immune therapy response. This suggests that assessing immune therapy response in ESCC patients through ITGB2 is feasible. The higher the expression of ITGB2 in patients, the greater the likelihood of benefiting from immune therapy. Additionally, we discovered that ITGB2 is significantly positively correlated with 10 macrophage targets that have entered clinical trials. This indicates that the higher the expression of ITGB2, the greater the feasibility of targeting macrophages. Furthermore, we identified three miRNAs associated with abnormal expression of ITGB2, providing reference for further exploration of upstream molecules influencing ITGB2 expression.

In summary, this study, through comprehensive integration of multiple datasets and methodologies, first identified a positive correlation between macrophage infiltration and ITGB2 expression in ESCC. ITGB2 is overexpressed in ESCC and holds potential as a prognostic marker for ESCC. As ESCC progresses, ITGB2 expression increases in infiltrating macrophages within the ESCC microenvironment. We also proposed for the first time that ITGB2 expression within macrophages increases as they evolve towards a tumor-promoting phenotype. We found that assessing immune therapy response in ESCC patients through ITGB2 is feasible, and higher expression of ITGB2 correlates with increased feasibility of targeting macrophages. Additionally, we identified three miRNAs associated with abnormal ITGB2 expression, providing reference for further exploration of ITGB2’s upstream molecular interactions.

There are some limitations in our study. Further validation through cell experiments is still necessary for our novel finding that the expression of ITGB2 increases as macrophages within ESCC shift towards a pro-tumor phenotype. Additionally, due to the specificity and complexity of the tumor microenvironment and tumor-associated macrophages, it will be necessary to combine emerging technologies such as spatial transcriptomics, high-dimensional flow cytometry, and multiplex immunohistochemistry in order to gain a more comprehensive understanding of the various subtypes and functional changes of tumor-associated macrophages in the tumor microenvironment of ESCC, as well as the more specific role of ITGB2 in this context, to better guide further basic and clinical research in the future.

## Data Availability

All raw data and code are available upon request from the corresponding author.
https://xena.ucsc.edu/
https://www.cancer.gov/ccg/research/genome-sequencing/tcga
https://www.ncbi.nlm.nih.gov/geo/

## AUTHOR CONTRIBUTIONS

Tao Huang was responsible for study design, data analysis, and manuscript writing. Longqian Wei and Jun Liu collected clinical samples, conducted WB and IHC experiments, and contributed to manuscript writing. Huafu Zhou managed the project, reviewed and revised the manuscript.

## ACKNOWLEDGMENTS

None.

## DATA AVAILABILITY STATEMENT

The publicly available data can be accessed through the respective website. Processed data and code can be obtained from the corresponding author upon request.

## ETHICS STATEMENT

This study was approved by the Ethics Committee of the First Affiliated Hospital of Guangxi Medical University, and informed consent was obtained from each patient.

